# Prioritising Hospital Complaints: An Innovative Tool Using Large Language Model-Assisted Content Analysis and Machine Learning Algorithms

**DOI:** 10.1101/2025.06.07.25329193

**Authors:** Muhammad Hafiz Sulaiman, Nora Muda, Fatimah Abdul Razak

## Abstract

**Background:** In clinical settings, patients often express dissatisfaction through narrative speech or written text. However, most complaints management systems still rely on manual review or rule-based methods that fail to capture the severity or urgency of complaints. This leads to inconsistent triage, delayed resolution and missed opportunities for systemic improvement. A novel model leveraging large language model-assisted content analysis (LACA) and machine learning (ML) can transform subjective narratives into standardized, machine-readable severity scores, facilitating the prioritisation of complaints.

**Objective:** This study aims to (1) determine the precision, recall fscore and accuracy of the proposed predictive models used to classify comments into low-alert and high-alert comment, (2) determine the construct validity and internal consistency (Cronbach’s α) of the themes found in LACA conducted on hospital web-based review data, (3) determine the predictors of low-alert and high-alert comments and their ability to change the log-odds of the outcome in logistic regression, and (4) to measure the robustness of the explanatory model measured by pseudo-R^2^.

**Methodology:** LACA was performed using a set of thematic codes to generate an independent variable dataset (*x*), with a scale of 0: not an issue, 1: a small issue, 2: a moderate issue, 3: a serious issue, and 4: an extremely serious issue. The independent variables (*x*) and the dependent variable (*y*, representing the review rating) were then split into training and testing sets to build predictive ML models. Grid search was used to determine the optimal combination of hyperparameters. The performance of the predictive and explanatory models was evaluated.

**Results:** ML classification was able to produce f1-score of 0.88 - 0.94 and accuracy of 0.92 for LR model; and f1-score of 0.87 - 0.94 and accuracy of 0.92 for ANN model The behaviour of predictive models was successfully explained by the explanatory model: Six (6) themes were determined with cumulative explained variance (CEV) of 0.74 and average Cronbach’s α of 0.86. LR shows significance on 5 themes with pseudo-R^2^ of 0.55.

**Conclusion:** This study demonstrates that a data pipeline utilizing LACA and ML algorithms shows excellent performance in classifying patient comments in a hospital setting. All effectiveness parameters including CEV, Cronbach’s α, precision, recall, f1-score, and accuracy indicate strong performance in differentiating high-alert from low-alert comments.

## Introduction

### Qualitative Nature of Patient Feedback and Complaints

#### United Kingdom

Evidence shows that hospital feedback channels overwhelmingly emphasize narrative (qualitative) input rather than just numbers. In the UK NHS, official guidance for the Friends & Family Test (FFT) requires at least one open-ended question for free-text comments. Indeed, the FFT manual explicitly notes that “free-text feedback can be the most important element” for driving improvement [1]. National patient surveys similarly rely on open comments. For example, the 2007 NHS Inpatient Survey included an “other comments” section – nearly 60% of respondents used it to write at least one comment [2]. Likewise, research on formal complaints finds that patients’ grievances are submitted as unsolicited written narratives. One systematic review noted that even the official Mid-Staffordshire NHS inquiry found its most important lessons in “unsolicited written patient complaints,” which revealed areas of poor care missed by structured metrics [3].

#### United States

According to Bufferne et al. (2023), in the US, patient satisfaction programs also collect large volumes of qualitative feedback. For instance, Press Ganey – a major survey provider – collects not only numeric scores but “a large number of free-text patient comments” from its surveys. One study analyzed nearly 65,998 open-ended comments provided by 48,592 hospitalized patients in a large Midwestern hospital system. The authors found that these comments captured issues not measured by the closed-ended survey questions, indicating that rich narrative feedback was a primary source of insight. In short, US hospital feedback systems similarly generate far more descriptive text (patient stories, concerns, suggestions) than simple ratings – and published analyses emphasize using NLP on these comments to inform improvement [4]

#### European Systems

Bufferne et al. (2023) also mentioned that European health systems follow the same pattern. France’s national patient survey (e-Satis) includes a “commentaires libres” section at the end. A French study found that 63.7% of respondents left at least one free-text comment. These narrative remarks were found to yield actionable information complementary to formal complaints. (Similarly, multi-country reviews note that structured experience surveys across Europe typically append open-ended questions, and countries like the UK and France require analysis of free-text feedback alongside quantitative scores.

#### Malaysia

Malaysia’s Public Complaint Management System (SISPAA) primarily collects qualitative, freetext feedback from the public, including in healthcare settings. SISPAA allows users to submit various forms of feedback, such as complaints, suggestions, compliments, inquiries, or comments, through an online platform. These submissions are typically unstructured and narrative in nature, rather than being confined to predefined rating scales or multiple-choice formats. [5] A study analysing complaints submitted via SISPAA to the Lembah Pantai Health Office between 2016 and 2018 found that the system effectively captured detailed, narrative complaints. The analysis focused on the content of these free-text submissions, highlighting the qualitative nature of the data collected. [6]

The prevalence of qualitative data in SISPAA presents both opportunities and challenges. While narrative feedback provides rich insights into patient experiences, the unstructured nature of the data can make systematic analysis and prioritization more complex. This underscores the potential value of integrating tools like Large Language Model-Assisted Content Analysis (LACA) and Machine Learning (ML) algorithms to process and interpret the data efficiently, facilitating more effective complaint management and service improvements.

#### Limitations of Manual and Rule-Based Complaint Handling

A systematic review of 59 studies revealed critical limitations in how healthcare complaints are analysed. Many existing tools lack a standardized taxonomy, fail to distinguish between different stages of care, and do not assess complaint severity. This inconsistency hampers the ability to accurately interpret and act upon complaints. [7] A study analysing 1,481 medical complaints in a tertiary hospital in Fujian Province highlighted that complaint data were collected and organized manually. The absence of a standardized management information system led to subjective judgments in classifying and resolving complaints, potentially overlooking critical issues. [8]

Traditional complaint categorization methods often focus on the type of issue rather than the harm caused. For instance, the Ombudsman Office at a large academic medical centre developed a Severity Scale to numerically classify complaints based on the harm caused, addressing the gap in existing frameworks that miss the opportunity to prioritize complaints based on severity. [9] Manual and rule-based methods may not effectively identify systemic problems. The Healthcare Complaints Analysis Tool (HCAT) was developed to provide a more reliable method for coding and analyzing healthcare complaints, enabling the identification of patterns and severity levels that traditional methods might miss. (Gillespie et al., 2016)

Quality improvement activities in hospitals rely heavily on feedback from patients and families to enhance care and ensure satisfaction, which is essential for patient retention and the financial viability of healthcare providers [10][11]. Traditional feedback methods, such as SERVQUAL surveys, have been used in Malaysian healthcare [12][13][14] but these can be time-consuming and limited in scope, leading to biased results due to small sample sizes and privacy concerns [15][16][17].

In contrast, web-based reviews provide a spatially unrestricted platform for patients to share their experiences at any stage of their hospital journey, offering a larger and more diverse set of feedback for analysis. [18] noted that web-based reviews cover a broader range of domains than traditional surveys like HCAHPS, which have fixed questions that may not capture evolving patient needs.

[19] emphasized the need for healthcare organizations to adapt to the fourth industrial revolution by leveraging web-based reviews to gain insights into patient desires and values, recommending the use of supervised machine learning to categorize these reviews into established SERVQUAL domains, thereby improving quality monitoring in hospitals.

#### LLM-Assisted Content Analysis

Thematic analysis is a qualitative research method that identifies, analyses, and reports patterns or themes within data. Originally, researchers begin by immersing themselves in the data, reading and reviewing it multiple times to gain a deep understanding. They then generate initial codes by tagging relevant sections with short labels that capture key aspects, which are organized into potential themes that reflect significant features related to the research question. After reviewing and refining these themes to ensure they accurately represent the data and fit cohesively, each theme is clearly defined and named. The final step involves writing up the findings, providing a comprehensive interpretation that weaves together the themes for insightful conclusions. Valued for its flexibility, thematic analysis effectively uncovers patterns within complex qualitative data, with much of the methodology in this study based on [20].

Original methods for analyzing text data from surveys and reviews face significant challenges, including time-consuming manual processes and resource-intensive efforts [21]. The vast volume and unstructured nature of textual data available online further complicate the extraction of actionable insights using conventional methodologies, prompting a shift towards automated content analysis [22]. The emergence of Large Language Models (LLMs), such as the gpt-4o-mini model, provides healthcare institutions with powerful tools to navigate and extract valuable insights from this data.

Text mining in big data analytics is gaining recognition for its ability to harness unstructured textual data, revealing significant patterns and correlations [23]. LLMs are sophisticated AI models trained on extensive text corpora, enabling them to understand and generate human-like language with remarkable fluency [24]. Their architecture, which incorporates attention mechanisms and feedforward neural networks, enhances functionality significantly [25]. With advanced natural language processing techniques, LLMs excel in text summarization, thematic analysis [26], and sentiment analysis [27], making them well-suited for qualitative data analysis in healthcare.

Recent studies have explored the impact of LLMs on drug discovery [28], medical note extraction [29], diagnosis-related group prediction [30], and diagnostics [31], highlighting opportunities for LLMs in clinical decision-making systems. In qualitative research, GPT’s context-awareness also allows it to produce direct textual descriptions, making it advantageous over Latent Dirichlet Allocation topic classification [32]. The advancement of LLMs aligns with global trends toward AI in healthcare, as noted by the World Health Organization [33], which emphasized the importance of AI and big data for improved healthcare delivery. Similarly, the Ministry of Health Malaysia prioritizes digitalization and AI in the country’s health reform agenda [34].

LLM-Assisted Content Analysis (LACA) refers to the use of Large Language Models (LLMs) to enhance qualitative content analysis. Researchers input qualitative data, such as text from webbased reviews, interviews or other documents, into an LLM, which processes and summarizes the content to provide an initial understanding. The LLM assists in coding and categorizing the text by suggesting themes and patterns through advanced natural language processing, thereby facilitating the identification and organization of key topics and underlying themes more efficiently [35]. Awais has demonstrated how to prompt GPT to produce a scalar data type and convert it into a binary data type. His study has shown the advantage of using scalar data type - it enables the researcher to optimize thematic analysis outcome (measured by human - LLM coders agreement) by manipulating threshold for scalar to binary data type conversion.

Our research questions are: (1) do the proposed predictive models perform well in classifying comments into high-alert and low-alert comments? (2) does LACA conducted on hospital webbased reviews produce good construct validity and internal consistency? (3) can we explain the behaviour of our predictive models using an explanatory model? (4) How strong is the proposed explanatory model in explaining the behaviour of the proposed predictive models? The purposes of this study are specifically to (1) determine the precision, recall f-score and accuracy of the proposed predictive models used to classify comments into low-alert and high-alert comment, (2) determine the construct validity and internal consistency (Cronbach’s α) of the themes found in LACA conducted on hospital web-based review data, (3) to determine the predictors of low-alert and high-alert comments and their ability to change the log-odds of the outcome in logistic regression, and (4) to measure the robustness of the explanatory model measured by pseudo-R^2^.

For these purposes, we developed three hypotheses: (1) the precision, recall, f-score and accuracy of predictive models using Artificial Neural Network (ANN) is higher than predictive models using Logistic Regression (LR), (2) the construct validity and internal consistency of the themes found in LACA shows high cumulative explained variance for factors with eigenvalues > 1 and high Cronbach’s α, (3) certain themes such as communication and professionalism have more influence than other themes in predicting the classification, and (4) the explanatory model has good to excellent pseudo-R^2^ indicating strong model fit in explaining the predictive model.

## Methodology

### Data collection and fake review exclusion

We use the data from web-based reviews that have hospital quality issues. No human participants were prospectively recruited for this study. All data were retrospectively collected from publicly accessible online reviews on 31^st^ December 2023. The data consist of one year of online reviews, from 1^st^ January 2023 to 31^st^ December 2023 and consist of 1,279 data points in text formats and already been filtered to remove fake reviews by using an ML filtering algorithm developed based on studies by [36][37][38][39]. The filtering algorithm involves NLP pre-processing, transforming into TF-IDF vector and training SVM model to classify fake versus real reviews using yelp.com data. Our data also excluded positive comments since [40] showed that bad reviews are the most meaningful to help readers make decisions about a hospital and to limit numbers of variables.

### GPT and Human Coding

A random 200 reviews were selected. gpt-4o-mini model API was tasked to use our codebook as a guide to code the 200 reviews. A human researcher was also tasked to code the same 200 reviews. Both GPT and human coders were instructed to code each review. The thematic coding is done by iterating each item in the codebook and scale the level of seriousness of the issues inside each webbased review. Scale 0 = no issue, scale 1 = small issue; scale 2 = moderate issue, scale 3 = serious issue; and scale 4 - extremely serious issue (refer **Appendix 1**). Since there are *n*_*i*_ items or codes in the codebook, the number of API calls will be 200*n*_*i*_. The human coder was only required to do the scale method, and the data was converted to binary for evaluating inter-rater reliability between human coder and GPT coder across the 200 reviews. Cohen’s Kappa, a statistical measure that accounts for chance agreement, is used for this purpose. The process of coding (described above) can then be continued solely by gpt-4o-mini model to the rest of our data.

### Predictive and Explanatory Model Development

To meet the dual objectives of this study (maximizing predictive performance and enhancing interpretability), we developed two separate machine learning pipelines, a predictive model and an explanatory model. Each model was constructed with a specific analytical purpose in mind, allowing us to balance data-driven performance with conceptual clarity.

The predictive model was designed to accurately classify patient comments into high-alert (low rating) and low-alert (high rating) categories. For this purpose, all original variables were directly used as inputs to the machine learning algorithms, without undergoing dimensionality reduction. This approach allowed us to preserve the full granularity of the dataset, enabling the model to detect subtle patterns and interactions that might be lost during abstraction. Modern machine learning algorithms, such as artificial neural networks (ANN) and logistic regression (LR), can effectively handle high-dimensional data. By retaining the complete variable set, the predictive model prioritized classification performance, which is essential for real-time complaint triage applications.

In contrast, the explanatory model was constructed to provide theoretical insight into the underlying drivers of patient dissatisfaction. This model utilized factor analysis to reduce the 41 observed variables into a smaller number of latent constructs, representing key themes such as communication, access, and service quality. These factor scores were then used in a logistic regression model to explain their association with high-alert classifications. This pathway enhances interpretability by mapping complex data into meaningful, conceptually aligned dimensions. Additionally, the use of factor scores mitigates issues of multicollinearity common in logistic regression, improving the model’s statistical robustness and stability.

Separating the predictive and explanatory models was necessary due to the trade-offs between performance and interpretability. While a factor-based model improves transparency and communication of findings, it may reduce classification accuracy due to abstraction. Conversely, a raw-variable model offers better predictive power but at the cost of complexity and reduced explanatory clarity. By employing both approaches, this study benefits from the strengths of each: high-performance classification for practical deployment and interpretable modeling for theoretical understanding and decision-making.

### Predictive Model Development

#### Data preprocessing of *y* Variable

Original *y* variable is online rating (with lowest rating = 1; and highest rating = 5). The *y* variable was non-normally distributed i.e. 72% of the *y* is distributed in extreme ends i.e. rating 1 (62%) and rating 5 (10%), while the rest in rating 2 (11%), 3 (8%) and 4 (9%). This extreme bias in rating was explained by Roh et al. (2021). The polarity suggests that it is wise for such data to be dichotomized to simplify analysis and interpretation. Training on original ordinal data leads to skewed model behaviour and difficulty capturing patterns in the minority classes (2,3 and 4). Binary classification is chosen because it is easier to interpret, more actionable (e.g flagging ‘high aler’ complaints), and better suited for priority triaging models. For such a reason, *y* variable was transformed by grouping rating 1 and 2 (73% of total data) as low rating and rating 4 and 5 (19% of total data) as high rating. Rating 3 and its *x* rows (8% of total data) were filtered out from the dataset since the total number is low and the rating is deemed neutral, thus removing it doesn’t affect predictability of the outcome.

#### Data Preprocessing of *x* Variables

Usually, data preprocessing involves addressing inconsistencies in scale, outliers, or skewed distributions that can bias model performance [41]. However, in this study, all thematic codes were generated using a consistent ordinal rating scale ranging from 0 (no issue) to 4 (extremely serious issue). This uniformity in measurement across variables ensured that each feature contributed proportionally to the learning process without dominating or being suppressed due to scale differences. Additionally, an exploratory analysis showed no presence of extreme outliers or heavy skewness that would necessitate corrective transformation. As a result, advanced scaling techniques such as robust scaling, log transformation, or quantile normalization were deemed unnecessary [^42^]. This consistency in feature representation not only simplified the preprocessing pipeline but also reduced the risk of introducing noise in data.

#### Splitting Data into Train and Test Datasets

To ensure a reliable evaluation of model performance, the dataset was divided into training and testing subsets using stratified sampling. This technique preserves the original distribution of class labels across both sets, which is particularly important given the imbalanced nature of the target variable, specifically, the underrepresentation of the high rating class [43]. Stratified splitting helps prevent biased performance estimates and ensures that the model is exposed to all classes during both training and evaluation. The typical split used was 80% for training and 20% for testing. [44]

#### Balancing Training Data

Imbalanced training datasets can lead to models that are overly biased toward predicting the low rating, resulting in poor sensitivity and failure to detect high rating cases. Balancing the training data is crucial for ensuring that the model can accurately identify and classify issues in hospital reviews, especially when the minority class represents low-alert or high-rating comments that should be given less priority [45]. To mitigate this problem, random oversampling technique were employed in this study, which artificially increase the number of minority class (high rating) samples by replicating existing ones. This approach helps the model learn decision boundaries more effectively, improving recall and reducing false negatives [46].

We applied oversampling only to the training set after splitting to avoid information leakage and ensure that the evaluation on the test set reflects real-world performance. If oversampling is performed before splitting, synthetic or duplicated samples of the minority class may appear in both the training and test sets. This overlap allows the model to “see” parts of the test data during training, leading to inflated performance metrics that don’t reflect real-world generalization. The purpose of the test set is to provide an unbiased evaluation of the model’s performance on unseen data. Oversampling before splitting compromises this by introducing artificially balanced data into the test set, which doesn’t represent the true distribution the model encounters in practice. [47]

#### ML Model Fitting

Following the balancing of the training dataset, two machine learning algorithms were developed and evaluated namely Logistic Regression (LR) and Artificial Neural Network (ANN). LR was selected due to its statistical interpretability, while ANN was chosen for its capacity to model complex nonlinear relationships. ANN Model training included hyperparameter optimization using grid search combined with 5-fold cross-validation (please refer ANN grid search hyperparameters below). The training dataset was partitioned into five equal subsets (folds). In each round, four folds were used to train the model and one fold was used for validation. This was repeated five times so that each fold served once as the validation set. The final performance score for each hyperparameter configuration was computed as the average f1-score across all five folds, which helped prevent overfitting and ensured robustness of model selection.

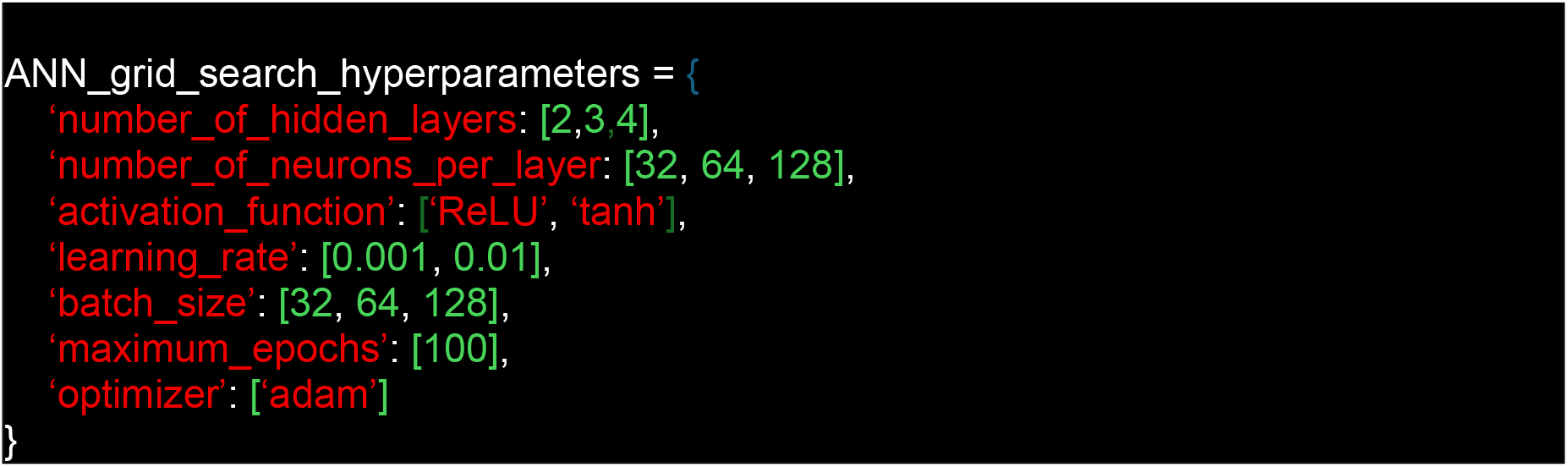

ANN were trained using mini-batch gradient descent, where model weights are updated after each batch rather than after the full dataset. Early stopping was employed by monitoring validation loss; training was halted if no improvement was observed over 10 consecutive epochs. The combination of grid search and cross-validation ensured that the models were tuned to generalize well beyond the training data, iteratively validate the model’s performance and reduce the risk of overfitting [48]. The best-performing hyperparameter sets were used to evaluate final model performance on the held-out test set. No hyperparameter tuning was conducted for logistic regression, as the model was fit using maximum likelihood estimation via the statsmodels Python package, which does not require regularization or solver specification.

#### ML Model Performance Measurement

Finally, model performance was assessed using a confusion matrix to calculate standard evaluation metrics: precision, recall, f1-score, and accuracy [49][50]. These metrics offer a comprehensive view of how well each model identified both positive (high alert) and negative (low alert) cases. Precision measures the proportion of true positives among predicted positives, while recall evaluates the ability to detect all actual positives. The f1-score balances these two, especially useful in imbalanced settings, and accuracy reflects the overall correctness of predictions. Together, these metrics ensure that the model is not only statistically sound but also clinically meaningful in flagging high-priority patient complaints.

### Explanatory Model Development

#### Themes Formation by Factor Analysis

To make the mechanisms influencing rating more explainable, *x* variables were reduced into a single-digit number using factor analysis. The use of factor analysis to reduce the number of thematic codes into factors (themes) is documented by [51]. We do a factor analysis as suggested by [52][53]. We decided to include / exclude factors based on cumulative explained variance, Cronbach α (George & Mallery, 2003)^54^ and qualitative assessment - content validity [55]. Full explanation on this can be found in our previously published paper [^56^]

#### Calculating Factor Score

*x* dataset underwent transformation into factor score using the transformer that has been trained using the train set before. The transformer calculates factor scores based on factor loadings of the 41 variables to reduce it to single-digit latent factors. Despite the factor scores not being directly interpretable, the factors act as secondary variables that can explain the relationship between these latent factors and rating.

#### Conducting Statistical Analysis

The factor scores were then fit into the statsmodel logit regression python module to determine which factors influencing online rating and the magnitude of their influence. Statistical models were then developed to summarize the mechanisms of determining online rating based on factors. Robustness (in terms of pseudo-R^2^) of the models developed were measured. Overall, the methodology involves a systematic approach to collect, integrate, analyse, and interpret data from web-based reviews to understand the factors influencing demand for private hospitals in Selangor. Advanced techniques like machine learning and natural language processing were used to filter out fake reviews and extract meaningful insights from large datasets

## Results

Of the 14,938 data collected, a total of 12,035 (81%) evaluations were accompanied by comments while 2,903 (19%) evaluations were not accompanied by comments. Among all reviews with comments, 1,121 (9.3%) reviews were fake and excluded from the data. There are 1,279 evaluations that have issues (negative sentiments) and 9,635 evaluations do not have issues (positive or neutral sentiments). The 1,279 evaluations give us the following analysis:

### Predictive Models

The best-performing ANN model consisted of an input layer with 41 features, followed by four hidden layers. The first hidden layer included 64 neurons with ReLU activation, the second had 128 neurons (ReLU), the third 64 neurons (ReLU), and the fourth 32 neurons (ReLU). The output layer had 2 neurons with a softmax activation function, supporting the one-hot encoding scheme used for binary classification. Best learning rate for our model is 0.001. The model was compiled using the Adam optimizer and trained with a categorical cross-entropy loss function. Training was performed over 20 epochs with a batch size of 32, and 20% of the training data was reserved for validation during training. This configuration provided the most favourable trade-off between model complexity and generalization performance, achieving high classification accuracy and f1scores on both training and test sets.

### Explanatory Model

#### Factor Analysis for Theme Formation

Scree plot shows 6 factors to be included in final themes (Figure 1). Full list of cumulative explained variance can be viewed in **Appendix 2**.

**Figure 1.**
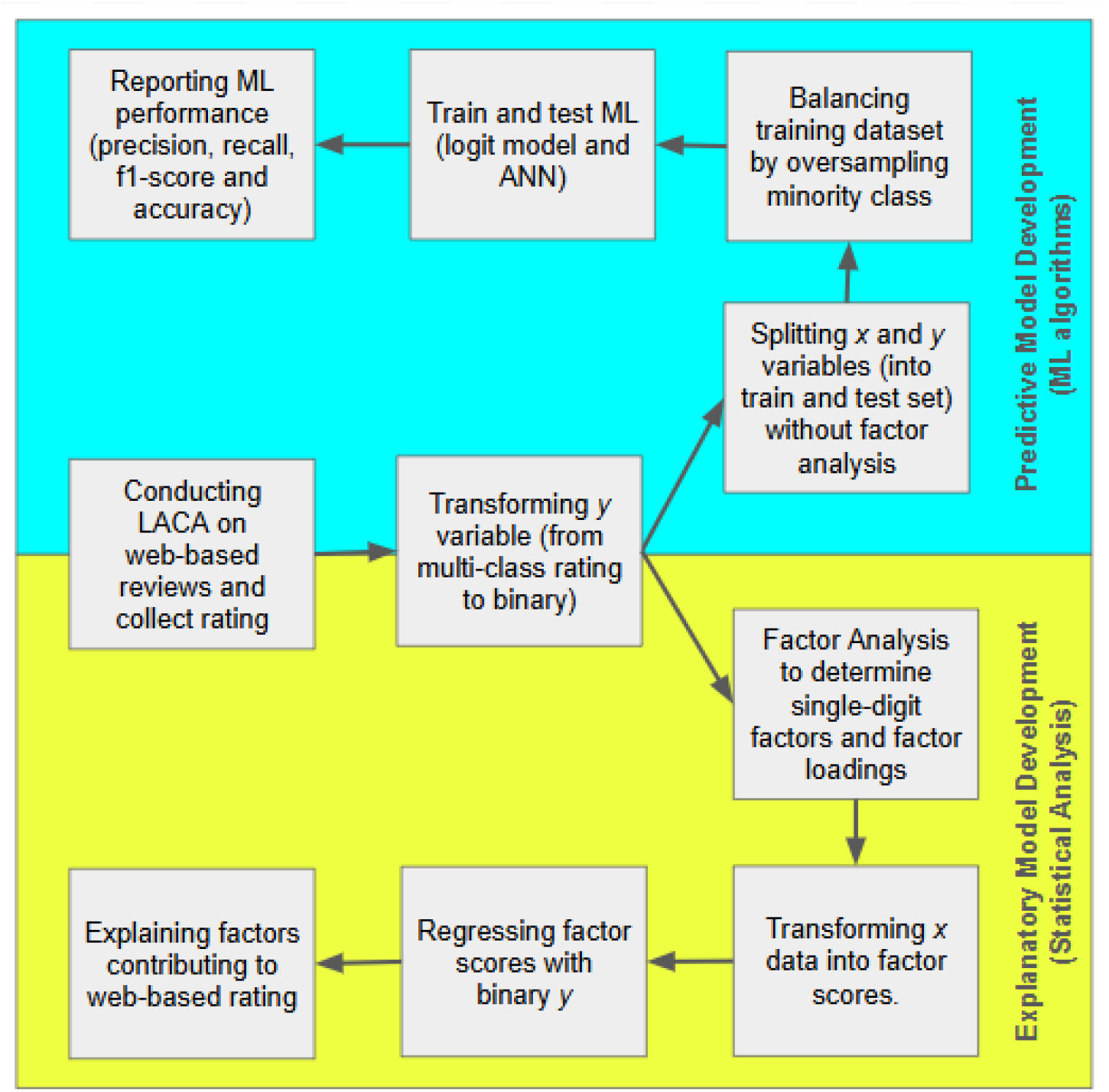
Flow of study methodology.

**Figure 6.**
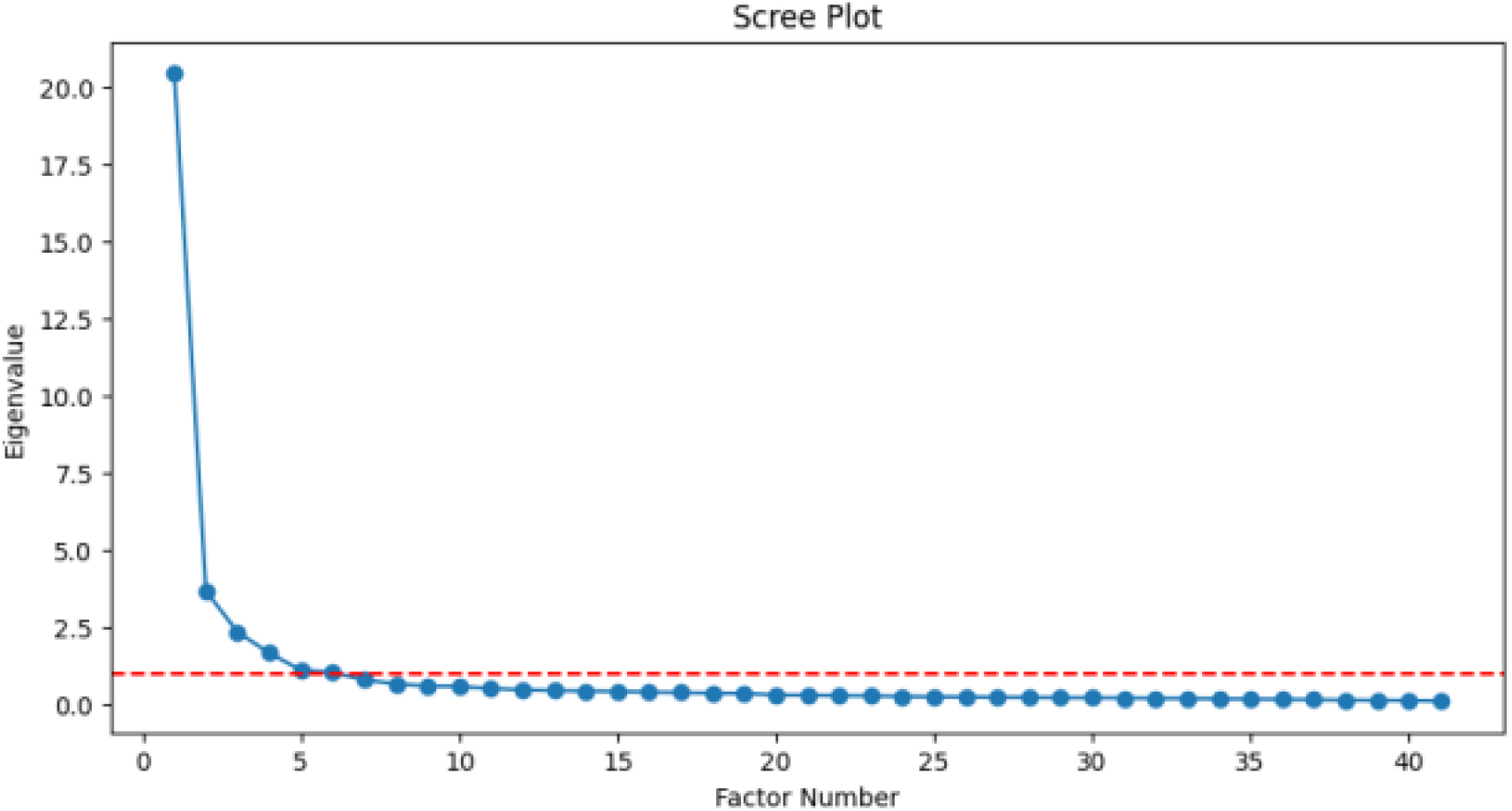
Scree plot.

## Discussion

The analysis of review content identified a substantial portion of evaluations (81%) accompanied by comments, with 19% lacking comments. Of the reviews with comments, 9.3% were deemed fake and excluded, leaving 9,594 evaluations without issues and 1,279 with issues. The use of gpt- 4o-mini model for coding these reviews showed a high inter-rater reliability, with an average Cohen’s Kappa score of 0.81, indicating strong agreement between human and AI coders. This high level of consistency supports the validity of the coding process, and the reliability of the insights derived from the data.

### Predictive Models Performance

Overall, the predictive models produce high precision, recall, f1-score and accuracy (Please refer **Table 1 and 2**). Prior to this study, we conducted an experiment using non-categorized ratings i.e. rating of 1, 2, 3, 4 and 5 instead of categorizing them into ‘high’ or ‘low’ rating. We found out that maintaining the *y* variable in its ordinal form brought difficulties in analysis and interpretation. As suggested by literature review, we also found imbalance in distribution of rating where rating 1 and 5, made the majority classes in the data. Our original data is imbalanced with 62% of ratings belonging to rating 1, 10% to rating 5, and 11%, 8% and 9% to ratings 2, 3, and 4 respectively.

**Table 1.** Confusion matrix for different machine learning algorithms.

| 1. Logistic Regression model |  |  |  |  |
| --- | --- | --- | --- | --- |
| Rating |  | Predicted |  |  |
|  |  | Low | High | Total |
| Actual | Low | 183 | 10 | 193 |
|  | High | 12 | 81 | 93 |
|  | Total | 195 | 91 | 286 |
| 2. Artificial Neural Network model |  |  |  |  |
| Rating |  | Predicted |  |  |
|  |  | Low | High | Total |
| Actual | Low | 185 | 8 | 193 |
|  | High | 15 | 78 | 93 |
|  | Total | 200 | 86 | 286 |

**Table 2.**
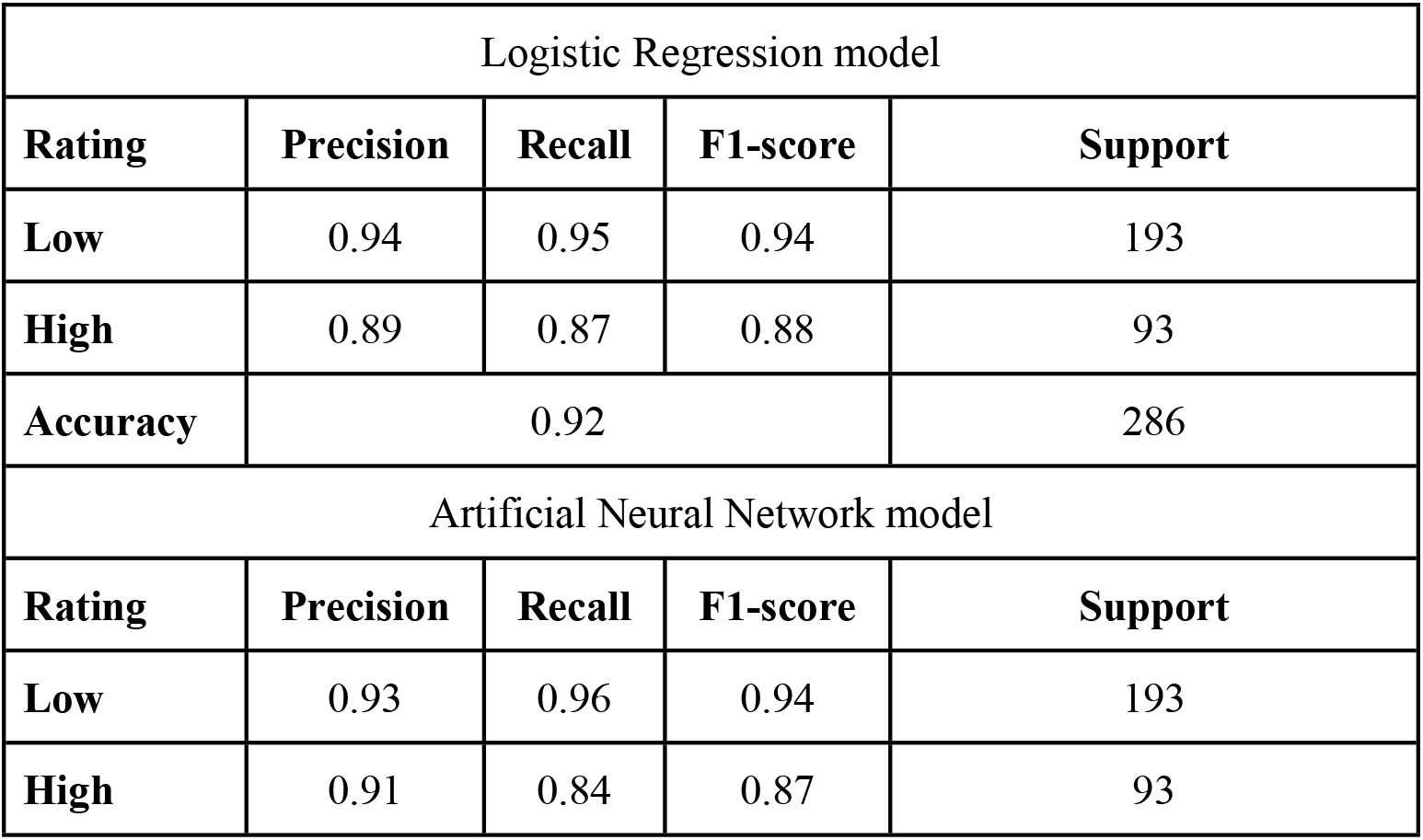
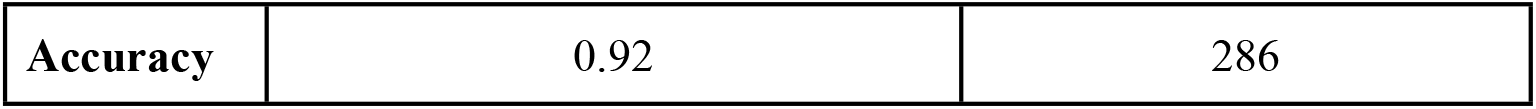
Classification report for different machine learning algorithms.

| Logistic Regression model |  |  |  |  |
| --- | --- | --- | --- | --- |
| Rating | Precision | Recall | F1-score | Support |
| Low | 0.94 | 0.95 | 0.94 | 193 |
| High | 0.89 | 0.87 | 0.88 | 93 |
| Accuracy | 0.92 |  |  | 286 |
| Artificial Neural Network model |  |  |  |  |
| Rating | Precision | Recall | F1-score | Support |
| Low | 0.93 | 0.96 | 0.94 | 193 |
| High | 0.91 | 0.84 | 0.87 | 93 |

|  |  |  |
| --- | --- | --- |
| Accuracy | 0.92 | 286 |

**Table 3.**
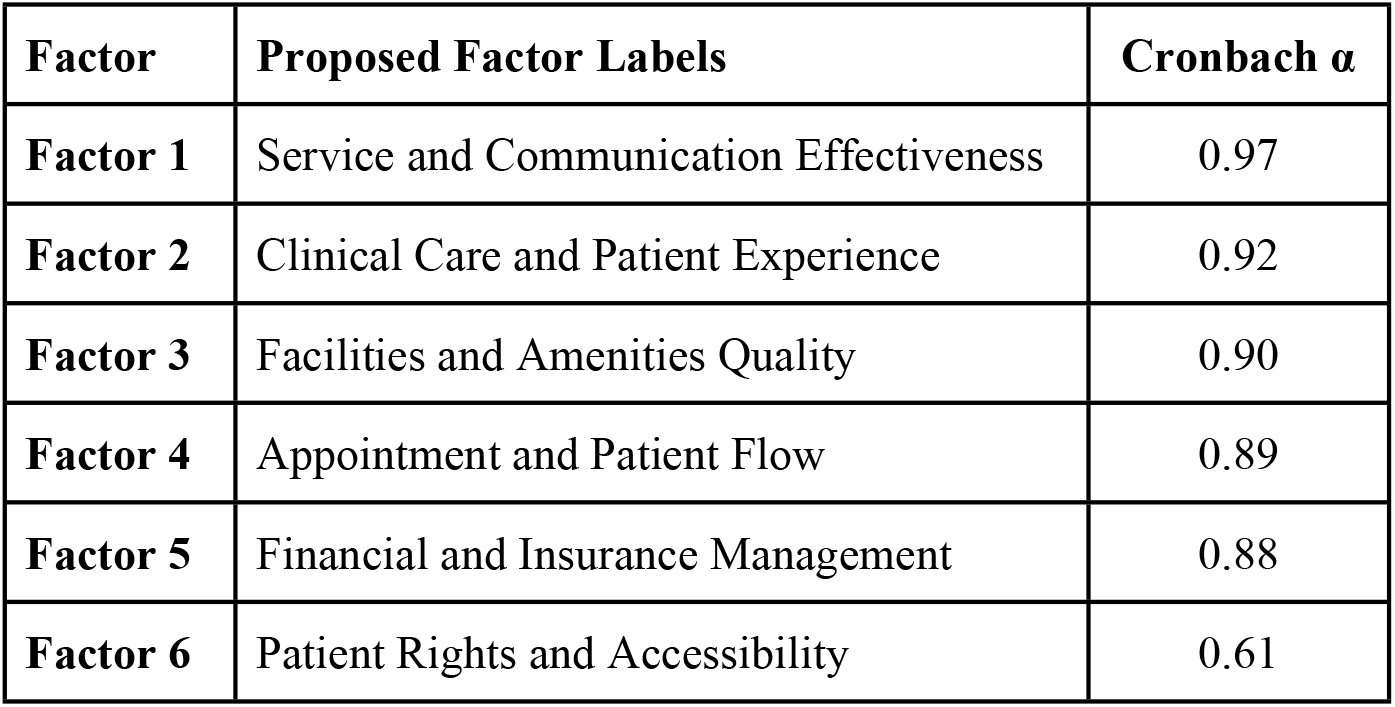
Retained factors and proposed labels.

**Table 4.** Logistic regression results.

| Factor | Coefficient | Std. Error | z | p value | CI (Min) | CI (Max) |
| --- | --- | --- | --- | --- | --- | --- |
| SCE | -2.0960 | 0.106 | -19.691 | < 0.001 | -2.305 | -1.887 |
| FAQ | -0.2849 | 0.078 | -3.670 | < 0.001 | -0.437 | -0.133 |
| APF | -0.4409 | 0.078 | -5.686 | < 0.001 | -0.593 | -0.289 |
| FIM | -0.2095 | 0.083 | -2.522 | < 0.001 | -0.372 | -0.047 |
| PRA | -0.5349 | 0.091 | -5.894 | < 0.001 | -0.713 | -0.357 |
| Pseudo-R<sup>2</sup> = 0.55 |  |  |  |  |  |  |

SCE - Service and Communication Effectiveness; FAQ - Facilities and Amenities Quality; APF - Appointment and Patient Flow; FIM - Financial and Insurance Management; PRA - Patient Rights and Accessibility.

**Table 5.** Summary of results for predictive models.

| $y$ data type | Model | f1-score | Accuracy |
| --- | --- | --- | --- |
| Binary | Logit | 0.88 & 0.94 | 0.92 |
|  | ANN | 0.87 & 0.94 | 0.92 |
| Multi-Class | Logit | 0.61 - 0.75 | 0.68 |
|  | ANN | 0.62 - 0.94 | 0.91 |

Even after oversampling of ratings 2, 3, and 4 (to balance the data), the precision, recall, and f1score remain low for ratings 2, 3, and 4 in all ML algorithms. This is due to significant overlap in terms of the variables distribution, making it difficult for the model to distinguish between these classes. If the differences between these ratings are subtle or not well represented by the features, the model struggles to accurately classify them, resulting in low performance for these classes. Despite balancing the class distribution, the model can also develop a bias toward predicting the majority classes (rating 1 and rating 5), especially because the minority classes (ratings 2, 3, and 4) do not have sufficient discriminative power. The model may predict the majority classes more often, which leads to low precision and recall for the minority classes.

Since the ML algorithms show high performances on extreme ends (rating 1 and 5), a reclassification of data made by transforming rating 1 and 2 to ‘low’ and rating 4 and 5 to ‘high’, and abandoning rating 3 (that has very low discriminative power) to produce a dichotomous category fit for logistic regression. The logistic regression shows a robust model with pseudo-R^2^ of 0.55. This shows that the transformation of the original ratings into a dichotomous ‘low’ and ‘high’ classification significantly enhances the predictive power of the model. By eliminating the less informative middle category (rating 3), the model can focus on the more discriminative extremes, leading to improved model performance.

ML algorithms for *y* binary classification were able to produce f1-score of 0.88 - 0.94 and accuracy of 0.92 for LR; and f1-score of 0.87 - 0.94 and accuracy of 0.92 for ANN model. It is also interesting to note that ANN algorithms produce lower accuracy as compared to logit similar to the finding in a study by [57]. Since our previous experiment using multi-class *y* variables shows better performance on ANN models as compared to LR model, this suggests that logit regression might be better at predicting binary *y* variable while ANN might be better at predicting multi-class *y* variable. Such assumption can be investigated further.

### Explanatory Model Performance

#### Factor Analysis

Factor analysis is a key component of our explanatory model. It identified six latent factors namely ‘Service and Communication Effectiveness’, ‘Clinical Care and Patient Experience’, ‘Facilities and Amenities Quality’, ‘Appointment and Patient Flow’, ‘Financial and Insurance Management’, and ‘Patient Rights and Accessibility’. The cumulative explained variance for the six variables is 0.74, which is strong for a social or community studies.

Cumulative explained variance (CEV) of 0.74, average Cronbach’s α of 0.86 and pseudo-R2 of 0.55 suggests that the model performs well at explaining factors influencing online rating. These results also indicate that our pipeline beginning from reclassification of *y* into binary data followed by the reduction of *x* variables using factor analysis, transformation of *x* variables into factor scores and logistic regression of the scores against the reclassified *y*, is effective in capturing meaningful patterns in the data, making the model a robust tool for explaining the factors influencing online rating. The log-odds formula to explain our model is derived from the Logistic Regression and is written as:

#### Formula of the Explanatory Model

Log(P(high)/P(low)) = -2.1*X*(Service and Communication Effectiveness)

-0.3*X*(Facilities and Amenities Quality)

-0.4*X*(Appointment and Patient Flow)

-0.2*X*(Financial and Insurance Management)

-0.5*X*(Patient Rights and Accessibility)

+0.02

Model’s pseudo-R^2^ = 0.55

In this model, the variable X represents a factor score, which is a latent (unobserved) continuous variable derived from the original ordinal ratings x (ranging from 0: ‘not an issue’ to 4: ‘an extremely severe issue’). The transformation from x to X is based on factor analysis that estimates how much each *x* contributes to a broader, underlying construct (e.g., “Service and Communication Effectiveness” or “Facilities and Amenities Quality”) etc. While x is directly observed and discrete, X captures a weighted, composite view of multiple ratings or themes. It adjusts for things like variance across items, correlation between different themes, and the relative importance of each rating level. As such, X becomes an abstract representation of the concept being measured, allowing for more precise modelling than raw ratings alone. This abstraction is essential in dimensionality reduction to interpret patient comments more meaningfully, especially when attempting to explain outcomes like high-alert complaints.

Each coefficient in the model represents the change in the log-odds of a patient comment being high alert (low rating) for a one-unit increase in that theme’s severity score, holding other variables constant. The largest negative coefficient (−2.1) is associated with Service and Communication Effectiveness, indicating that concerns in this area are the strongest predictor of low-rating comments, consistent with a finding reported by [58]. Patient Rights and Accessibility (−0.5) and Appointment and Patient Flow (−0.4) also meaningfully contribute to predicting low-rated comments, although their impact is less pronounced than communication issues [59]. The smallest coefficient is for Financial and Insurance Management (−0.2), suggesting that while still relevant, its influence on complaint severity is lower relative to other themes, which finding is supported by [60]. The intercept of +0.02 indicates a slight bias toward the low-rating classification when all predictors are zero, though this has minimal practical effect.

#### Explanatory Model Fit and Implications

The model achieves a pseudo-R2 value of 0.55, which suggests a strong explanatory power in the context of logistic regression, according to [61][62] that stated a McFadden’s pseudo R^2^ of 0.2 and 0.4 are often considered good. This means that over half the variance in the binary outcome (low vs high alert) is explained by the five thematic variables in the model. In the context of social science or healthcare models, this is considered a robust fit. This model highlights which themes are most associated with serious dissatisfaction or alert-worthy patient comments. Particularly, issues related to service quality and communication are strong drivers of negative perceptions, which may warrant prioritization in complaints management efforts. The model also demonstrates the potential of combining structured content analysis with statistical modelling to objectively triage qualitative feedback.

## Conclusion

Our analysis revealed clear and meaningful latent structures through factor analysis, yielding high cumulative explained variance (CEV = 0.74) and strong internal consistency (average Cronbach’s α = 0.86). These results confirm the reliability and conceptual soundness of the thematic coding process used in large language model-assisted content analysis (LACA). The machine learning models, particularly Logistic Regression (LR) and Artificial Neural Networks (ANN), demonstrated excellent performance in classifying patient comments into high-alert (low rating) and low-alert (high rating) categories. Both models achieved high precision, recall, f1-score (0.87– 0.94), and accuracy (0.92), validating the effectiveness of the proposed data processing pipeline. Logistic Regression showed strong explanatory power with a pseudo-R2 of 0.55, reinforcing the utility of transforming original ratings into a dichotomous classification. While challenges such as class imbalance existed in the beginning, the reclassification strategy enhanced model performance and interpretability. Overall, the integration of LACA with ML presents a powerful approach to healthcare review analysis, supporting more accurate complaints triage, deeper insight into patient satisfaction, and the development of scalable, robust predictive systems. Future studies could be conducted to evaluate the effectiveness of the proposed models in actual clinical settings.

## Data Availability

The data used to train ML, conduct factor analysis and build explanatory model can be found here: https://drive.google.com/file/d/1Pw1eRAzjOmm7-wcP1YtfBguqHPtnA8YR/view?usp=sharing Patient complaints is publicly available in google review pages. Some hospitals are sensitive about this data so we decided not to share it publicly because it mention names of hospitals and medical staff.

## Appendix 1

API call for giving a scale for each thematic codes on each web-based review.

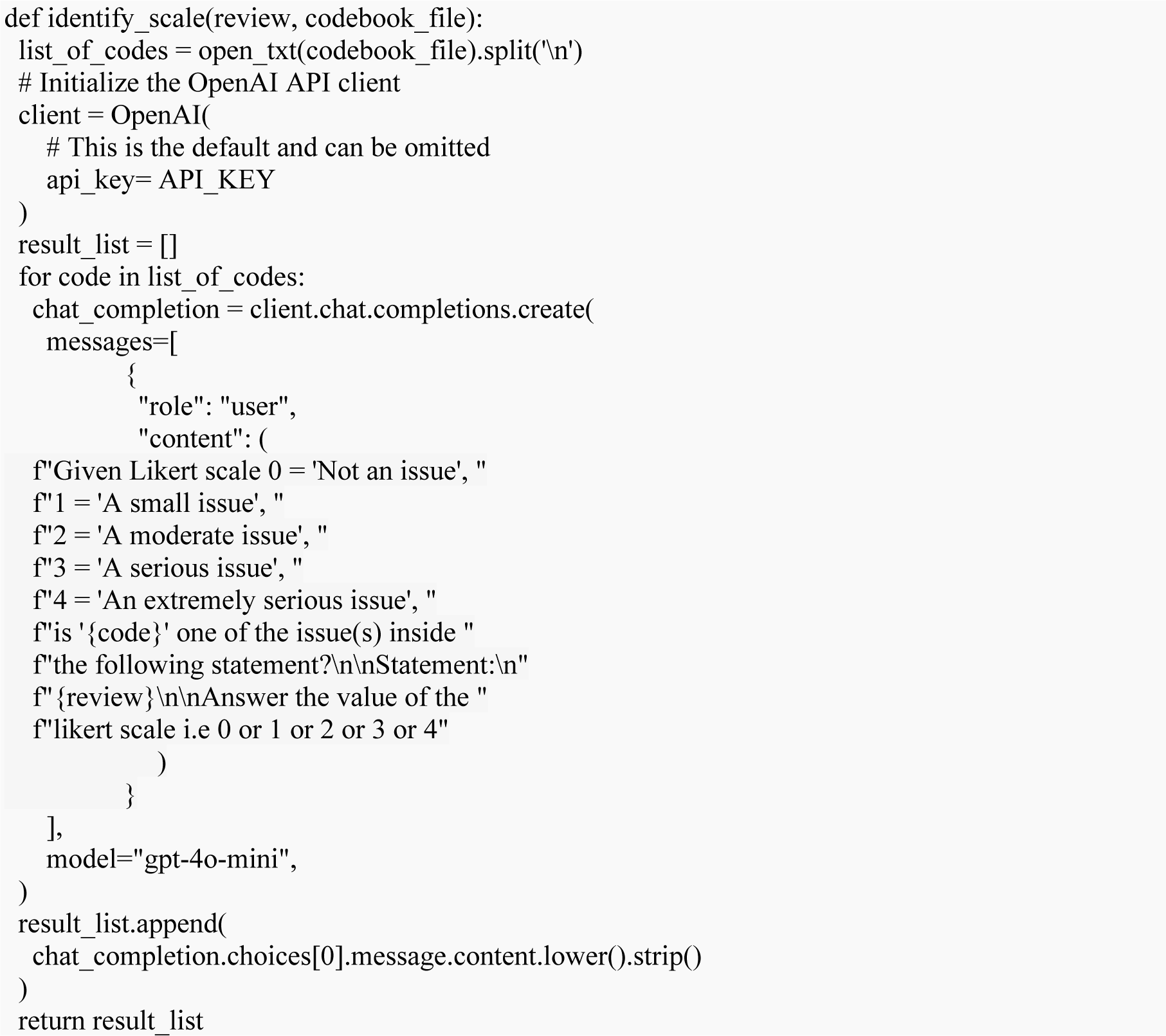

## Appendix 2

Cumulative Explained Values

| Cumulative Explained Values (F01 - F41) |
| --- |
| F01: 0.50, F02: 0.59, F03: 0.64, F04: 0.69, F05: 0.71, F06: 0.74, F07: 0.76, F08: 0.77, F09: 0.79, F10: 0.80, F11: 0.81, F12: 0.82, F13: 0.84, F14: 0.85, F15: 0.86, F16: 0.87, F17: 0.88, F18: 0.89, F19: 0.90, F20: 0.90, F21: 0.91, F22: 0.91, F23: 0.92, F24: 0.93, F25: 0.95, F26: 0.96, F27: 0.96, F28: 0.97, F29: 0.98, F30: 0.99, F31: 0.99, F32: 1.00, F33: 1.00, F34: 1.00, F35: 1.00, F36: 1.00, F37: 1.00, F38: 1.00, F39: 1.00, F40: 1.00, F41: 1.00. |

